# Balancing data quality and participant burden: A comparative analysis of abbreviated vs extended symptom diaries in the CanTreatCOVID trial

**DOI:** 10.64898/2026.01.23.26343617

**Authors:** Benita Hosseini, Dorsa Mohammadrezaei, Parsa Balalaie, Kawsika Sivayoganathan, Amanda Condon, Bruno R. da Costa, Peter Daley, Michelle Greiver, Peter Jüni, Todd C. Lee, Kerry McBrien, Emily G. McDonald, Srinivas Murthy, Peter Selby, Andrew D. Pinto

## Abstract

**Background:** Symptom diaries are widely used in acute respiratory infection trials to capture patient-reported symptom severity and recovery. Longer questionnaires may provide a more complete clinical picture but can increase participant burden and reduce adherence. Evidence directly comparing long and short formats within the same trial is limited.

**Objective:** To compare adherence, symptom trajectories, agreement between recovery measures, and predictive performance for recovery-related outcomes between a short and long symptom diary in an outpatient SARS-CoV-2 trial

**Methods:** This secondary analysis of the CanTreatCOVID trial compared a 9-item Abbreviated Diary and a 34-item FLU-PRO Plus Diary over 14 days in non-hospitalized participants with confirmed SARS-CoV-2 infection. Outcomes included diary initiation, completion, completion rate, compliance, symptom trajectories, agreement between recovery outcomes, and predictive performance. Analyses used logistic regression, generalized estimating equations, survival models, and predictive modelling.

**Results:** Of 712 participants, 638 used the Abbreviated Diary and 74 the FLU-PRO Plus Diary. Baseline characteristics were similar between groups. Diary type was not significantly associated with diary initiation or full 14-day compliance, whereas treatment assignment was associated with higher adherence (p < 0.0001). Completion rates were slightly higher in the Abbreviated group (68.1% vs. 64.4%), but differences were not statistically significant. Agreement between “feeling recovered” and “return to usual health” was strong to excellent (κ = 0.8371–0.8859), while agreement with “return to usual activities” was moderate (κ = 0.5273–0.6583). Predictive models performed well for both diaries (AUCs 0.87–0.94), with only marginal gains from including the extended FLU-PRO Plus items.

**Conclusion:** In this outpatient SARS-CoV-2 trial, abbreviated and extended symptom diaries produced comparable adherence, symptom trajectories, and predictive performance. Future research should extend follow-up beyond 14 days to capture longer-term patterns and test diary performance in more diverse and digitally underserved populations.

## Introduction

Symptom diaries are widely used in clinical trials for acute respiratory infections to capture patient-reported symptom severity and recovery over time.^1,2^ They provide longitudinal data that complement clinician assessments and enable early evaluation of treatment effects, particularly in community-based trials where in-person follow-up is limited.^2^ Diary formats vary considerably, and the design of these diaries must balance the need for comprehensive data with the burden placed on participants.^3^

Previous research has demonstrated that questionnaire length has a strong impact on adherence. Meta-analyses^4^ and studies in both clinical and non-clinical contexts have found that longer questionnaires are consistently associated with lower completion rates^5–7^, while shorter formats improve retention without introducing systematic bias in responses^4,5,7^. Although data quality may remain stable, longer tools increase the risk of fatigue^5^, and the benefits of added detail may be offset by reduced participant engagement over time.

In the context of symptom diaries, longer formats may provide a more complete clinical picture but can reduce adherence.^8,9^ Shorter diaries are easier to complete^10^ yet may omit relevant symptoms that could be important for understanding disease course or treatment effects.^9^ Several studies have evaluated the use of symptom diaries for SARS-CoV2 and other respiratory infections. The FLU-PRO Plus, a validated 34-item diary, captures a wide range of symptom domains and has been adapted for use in COVID-19 trials.^11^ It has shown strong validity and responsiveness, although long-term adherence data, especially in real-world settings, is limited^12^. In contrast, shorter symptom diaries with 8–10 core symptoms have been used in large adaptive platform trials such as PANORAMIC^13^ and PRINCIPLE^14,15^ in the United Kingdom. These abbreviated tools are more feasible for widespread implementation and can achieve higher retention but may miss less common or emerging symptoms,^13^ especially during the emergence of new variants or in specific subgroups.

The balance between data richness and feasibility is a well-recognized challenge in trial design. However, evidence directly comparing long and short diary formats within the same trial is limited, and little is known about how diary length affects data quality, recovery estimates, and predictive modeling of outcomes. The CanTreatCOVID trial^16^ offers a unique opportunity to address this gap. In this secondary analysis, we compared a 9-item Abbreviated Diary and the 34-item FLU-PRO Plus Diary used in the trial. Our objective was to examine adherence, symptom severity trajectories, and predictors of recovery across the two diary formats. The focus of this analysis is on differences between diary types rather than treatment effects, with the aim of informing future trial designs that require reliable symptom monitoring while minimizing participant burden.

## Methods

### Study design and population

This is a secondary analysis of data from the CanTreatCOVID trial, an open-label, individually randomized, multi-centre adaptive platform trial evaluating the effectiveness of therapeutics for non-hospitalized individuals with symptomatic SARS-CoV-2 infection in Canada. Eligibility criteria, recruitment strategies, and study procedures are described in detail elsewhere.^16^

Briefly, eligible participants had symptomatic SARS-CoV-2 infection confirmed by PCR or rapid antigen test within five days of symptom onset. The trial targeted individuals at higher risk for severe outcomes, including those aged ≥50 years and those aged 18–49 years with at least one underlying comorbidity (i.e. cardiovascular disease, diabetes, chronic respiratory illness). Participants were randomized to receive either an active treatment or usual care and were asked to complete a structured online symptom diary daily for 14 days following randomization (Supplementary File 1).

The Abbreviated Diary captured self-reported severity for 9 core symptoms using a 4-point Likert scale ranging from 0 (none) to 3 (major problem), a method validated in prior similar trials. ^13,17,18^ A randomly selected 10% of participants were assigned to the FLU-PRO Plus diary, which included the same 9 core symptoms plus 34 additional symptom questions covering multiple domains (i.e. systemic, nasal, throat, gastrointestinal, and chest symptoms; Supplementary File 2).

### Data collection

Participants were prompted to complete daily surveys via REDCap Cloud© through automated email reminders. For those without internet access or who preferred phone follow-up, research staff conducted surveys by phone. If diaries were incomplete for two or more consecutive days before day 7 or 14, staff followed up by email, phone, or text, making up to three contact attempts over three days before considering it missing data.

### Outcomes

The primary focus of this analysis was to compare adherence patterns and symptom reporting across the two diary types. Diary adherence outcomes included completion of at least one diary entry, completion of all 14 diary entries (compliance), average completion rate across the 14-day follow-up period, time to first missed diary entry (diary dropout), and daily response behavior over time. Symptoms-related outcomes included daily symptom severity trajectories, agreement between three binary recovery indicators (“feeling recovered,” “return to usual health,” and “return to usual activities”), predictors of recovery, and time to recovery. Outcomes were assessed using the nine core symptoms in both groups; in the FLU-PRO Plus group, extended analyses incorporated the full 34-symptom set.

### Statistical analysis

Descriptive statistics were used to summarize all outcomes by diary type and treatment group. For diary compliance, logistic regression models were used to assess the association between diary type or treatment assignment and the binary outcomes of completing at least one entry or completing all 14 entries. Linear regression was used to compare mean completion rates between groups. A multivariable linear regression model was fitted to examine associations between baseline characteristics and completion rate, adjusting for treatment group, age, sex, race, education and income level, housing status, employment status, health status, and baseline SARS-CoV-2 symptom severity. Differences in baseline characteristics between diary groups were evaluated using chi-square tests. Time to first missed diary entry was analyzed using Kaplan-Meier survival analysis, and the log-rank test was used to compare dropout curves between diary groups. A Cox proportional hazards model was then used to identify factors associated with dropout, with covariates including diary type, demographic characteristics, health status, and SARS-CoV-2 symptom severity. Daily response behavior over the 14-day period was modeled using a weighted generalized estimating equations (GEE) approach with inverse probability weighting to account for imbalance in diary group assignment, including diary type, treatment group, time, and their interactions, with an exchangeable correlation structure to account for repeated measures.

For symptom trajectory analysis, linear mixed-effects models were used to examine changes in daily symptom severity scores over the 14-day period. Fixed effects included day, treatment group, diary type, age, sex, and vaccination status, along with interaction terms between day, diary type, and treatment to assess differences in trajectories by group. Random intercepts for participant ID were included to account for repeated measures.

To evaluate the consistency of recovery measures, we assessed the agreement between the primary indicator of self-reported recovery (“Do you feel recovered today?”) and two related metrics: “Return to usual health” and “Return to usual activities.” Analyses were stratified by diary type and treatment group. Agreement was quantified using Cohen’s Kappa coefficient (κ), with values >0.80 interpreted as strong agreement and values between 0.6–0.8 as moderate agreement.

To evaluate the relationship between daily symptom burden and recovery outcomes, GEE models with an exchangeable correlation structure were fitted separately for the Abbreviated Diary and FLU-PRO Plus groups. Models were stratified by diary type and predicted three outcomes: feeling recovered, return to usual health, and return to usual activities. Each model included daily symptom reports and baseline characteristics and accounted for repeated measures across participant-days.

Cox proportional hazards models were used to identify baseline symptoms and demographic characteristics associated with time to self-reported recovery within the FLU-PRO Plus diary group. Two models were evaluated: one including only the nine most clinically prominent symptoms (common symptoms model) and another incorporating all 34 symptoms recorded in the diary (extended symptoms model). Both models adjusted for treatment group, age group, sex, and vaccination status.

Participants not reporting recovery within 14 days were censored. All statistical tests were two-sided, and p-values < 0.05 were considered statistically significant. Analyses were conducted using R (version 4.3.0) and SAS (version 9.4).

## Results

A total of 712 participants were included (638 Abbreviated Diary, 74 FLU-PRO Plus). Baseline characteristics were well balanced across groups, with no statistically significant differences in demographics, socioeconomic status, health status, or SARS-CoV-2 severity **(Table 1).**

**Table 1.** – Baseline characteristics of participants by diary type.

| **Variable** | **Group** | **Abbreviated N (%)** | **FLU-PRO Plus N (%)** | **p-value** |
| --- | --- | --- | --- | --- |
| Age | 18-49 | 163 (25.5%) | 18 (24.3%) | 0.685 |
|  | 50-65 | 333 (52.2%) | 42 (56.8%) |  |
|  | More than 65 | 141 (22.1%) | 14 (18.9%) |  |
| Sex | Female | 415 (65.0%) | 51 (68.9%) | 0.727 |
|  | Male | 195 (30.6%) | 21 (28.4%) |  |
| Race | Non-White | 138 (21.6%) | 15 (20.3%) | 1.000 |
|  | White | 499 (78.2%) | 59 (79.7%) |  |
| Education | High School or Less | 49 (7.7%) | 4 (5.4%) | 0.177 |
|  | University/College | 364 (57.1%) | 51 (68.9%) |  |
|  | Graduate | 198 (31.0%) | 17 (23.0%) |  |
| Housing | Renting/Other | 186 (29.2%) | 29 (39.2%) | 0.055 |
|  | Own Home | 451 (70.7%) | 45 (60.8%) |  |
| Employment | Non-Employed | 276 (43.3%) | 38 (51.4%) | 0.172 |
|  | Employed | 361 (56.6%) | 36 (48.6%) |  |
| Income | <60,000 | 135 (21.2%) | 18 (24.3%) | 0.100 |
|  | 60,000-100,000 | 142 (22.3%) | 23 (31.1%) |  |
|  | >100,000 | 329 (51.6%) | 29 (39.2%) |  |
| Chronic Disease | Yes | 474 (74.3%) | 58 (78.4%) | 0.665 |
|  | No | 137 (21.5%) | 14 (18.9%) |  |
| Self Reported<br>Health | Poor/Fair | 91 (14.3%) | 12 (16.2%) | 0.942 |
|  | Good | 202 (31.7%) | 22 (29.7%) |  |
|  | Very good | 235 (36.8%) | 27 (36.5%) |  |
|  | Excellent | 83 (13.0%) | 11 (14.9%) |  |
| SARS-CoV2<br>Severity | No symptoms/Mild | 165 (25.9%) | 19 (25.7%) | 0.828 |
|  | Moderate | 384 (60.2%) | 44 (59.5%) |  |
|  | Severe/Very severe | 62 (9.7%) | 9 (12.2%) |  |

### Diary completion and compliance analysis

Overall, 92.4% of participants completed at least one diary entry, with similar proportions in the Abbreviated (92.5%) and FLU-PRO Plus (91.9%) groups.

Diary type was not significantly associated with the likelihood of completing at least one entry (p>0.05). Completion rate, defined as the percentage of expected diary entries completed over 14 days, was slightly higher in the Abbreviated Diary group (71.4% Paxlovid, 63.3% Usual Care) than in the FLU-PRO Plus group (69.3% Paxlovid, 58.6% Usual Care), though this difference was not statistically significant (p = 0.239) **(Table 2)**.

**Table 2.**
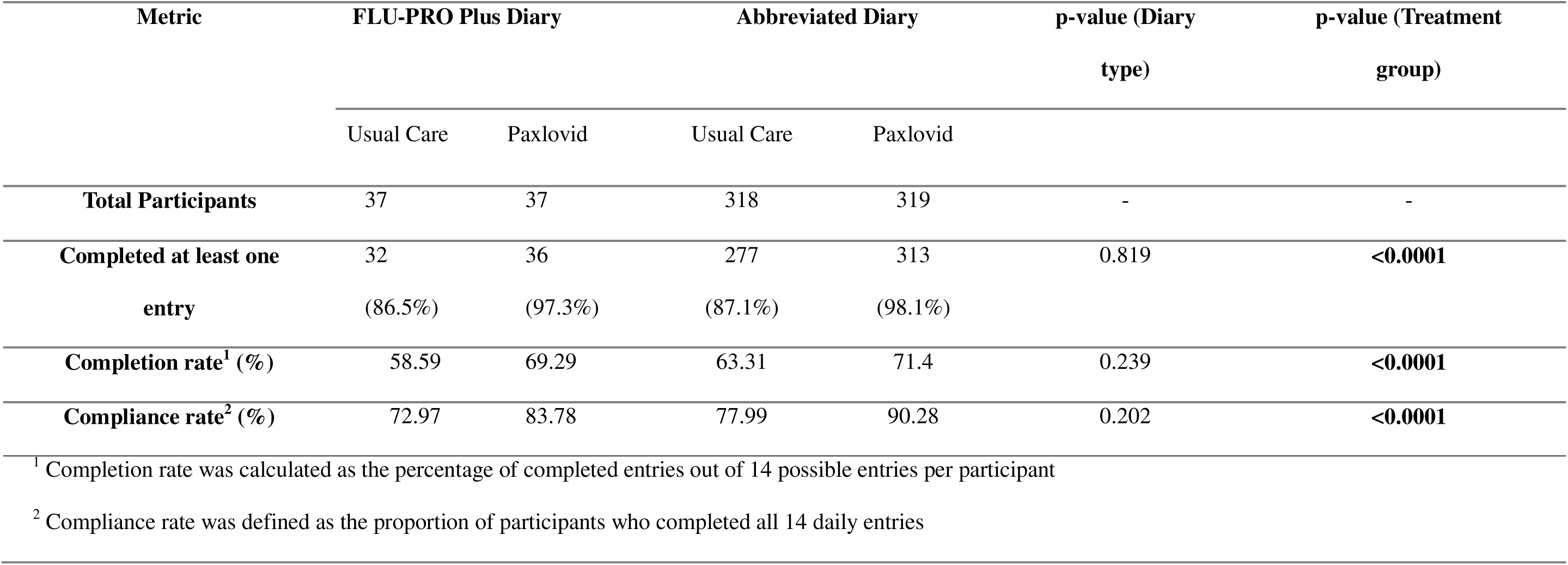
– Participant completion outcomes by diary type and treatment group.

| Metric | FLU-PRO Plus Diary |  | Abbreviated Diary |  | p-value (Diary type) | p-value (Treatment group) |
| --- | --- | --- | --- | --- | --- | --- |
|  | Usual Care | Paxlovid | Usual Care | Paxlovid |  |  |
| **Total Participants** | 37 | 37 | 318 | 319 | - | - |
| **Completed at least one entry** | 32<br>(86.5%) | 36<br>(97.3%) | 277<br>(87.1%) | 313<br>(98.1%) | 0.819 | <0.0001 |
| **Completion rate<sup>1</sup> (%)** | 58.59 | 69.29 | 63.31 | 71.4 | 0.239 | <0.0001 |
| **Compliance rate<sup>2</sup> (%)** | 72.97 | 83.78 | 77.99 | 90.28 | 0.202 | <0.0001 |

<sup>1</sup> Completion rate was calculated as the percentage of completed entries out of 14 possible entries per participant
<sup>2</sup> Compliance rate was defined as the proportion of participants who completed all 14 daily entries

Compliance, defined as completing all 14 diary entries, was also higher in the Abbreviated group (90.3% Paxlovid, 78.0% Usual Care) compared to the FLU-PRO Plus group (83.8% Paxlovid, 72.9% Usual Care), but the difference did not reach statistical significance (p = 0.202) **(Table 2)**.

Within each diary type, participants assigned to study treatment (nirmatrelvir/ritonavir (Paxlovid), administered two times per day for 5 days) were significantly more likely to initiate diary completion (p < 0.0001) and had higher overall completion rates and compliance rate compared with those receiving usual care (p < 0.0001) groups **(Table 2)**.

To account for group size imbalance between diary types, we performed a downsampling procedure by randomly selecting an equal number of participants from each group and re-running regression models as a sensitivity analysis. The downsampled analysis yielded almost consistent results, confirming that diary type was not a significant predictor of diary completion rate (p = 0.095), whereas treatment group remained a significant predictor, with higher completion rates observed in the Paxlovid group (p = 0.013).

In multivariable linear regression adjusting for baseline characteristics and treatment group, higher completion rates were observed among White participants (p = 0.002) and those assigned to Paxlovid (p = 0.011) **(Supplementary Table S1)**. No significant associations were found for age, sex, education, employment, income, chronic disease status, self-reported health, or SARS-CoV-2 severity. In weighted GEE models, Paxlovid was associated with higher odds of daily response (OR = 6.18, p = 0.003), whereas diary type was not (OR = 0.74, p = 0.507

Daily completion rates declined over time in both diary groups, with a steeper decline in the FLU-PRO Plus group **(Figure 1)**.

**Figure 1.**
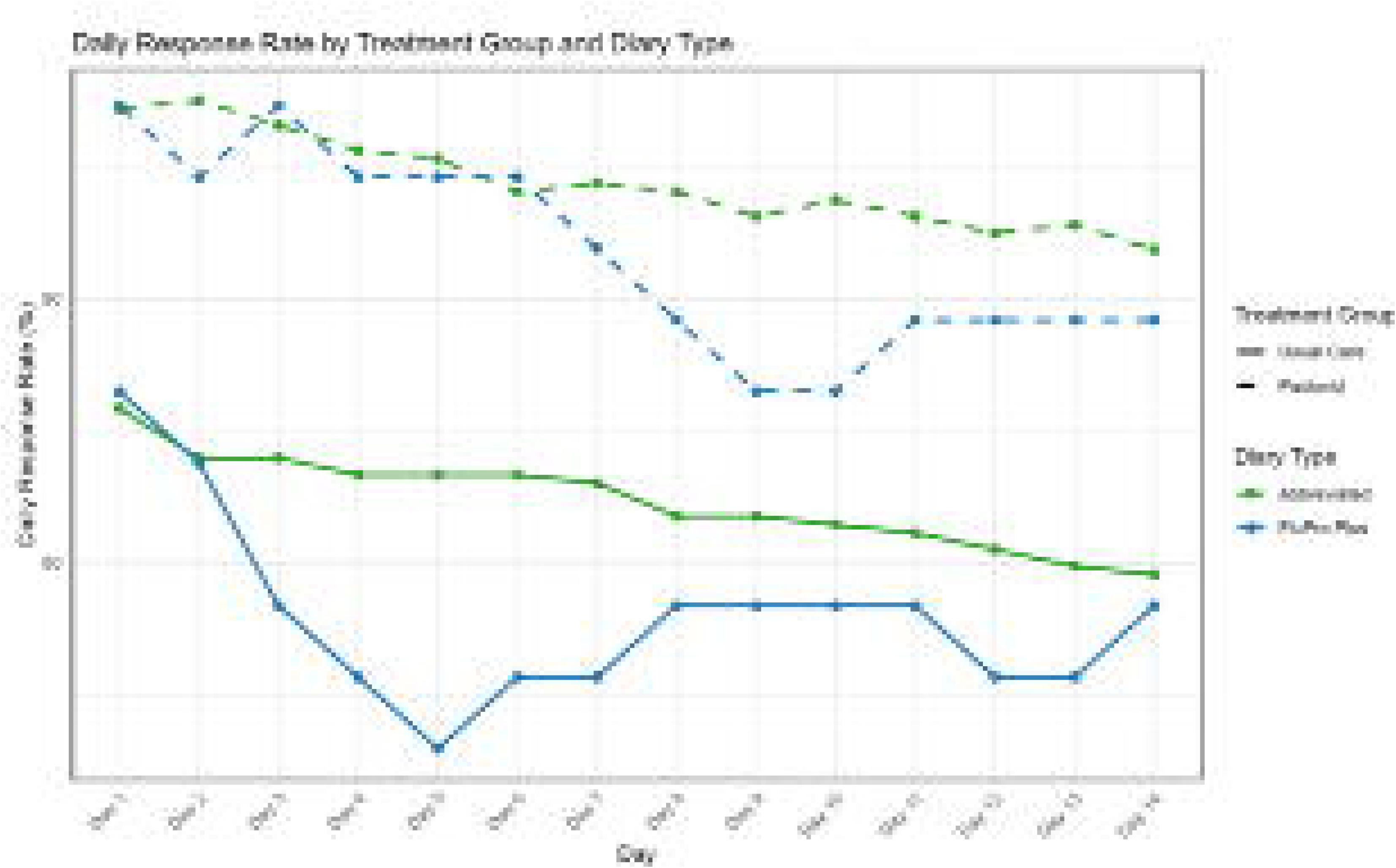
Daily response rate by treatment group and diary Type.

Kaplan–Meier analysis suggested earlier discontinuation in the FLU-PRO Plus group, although survival curves were not significantly different (log-rank p = 0.20) **(Figure 2)**.

**Figure 2.**
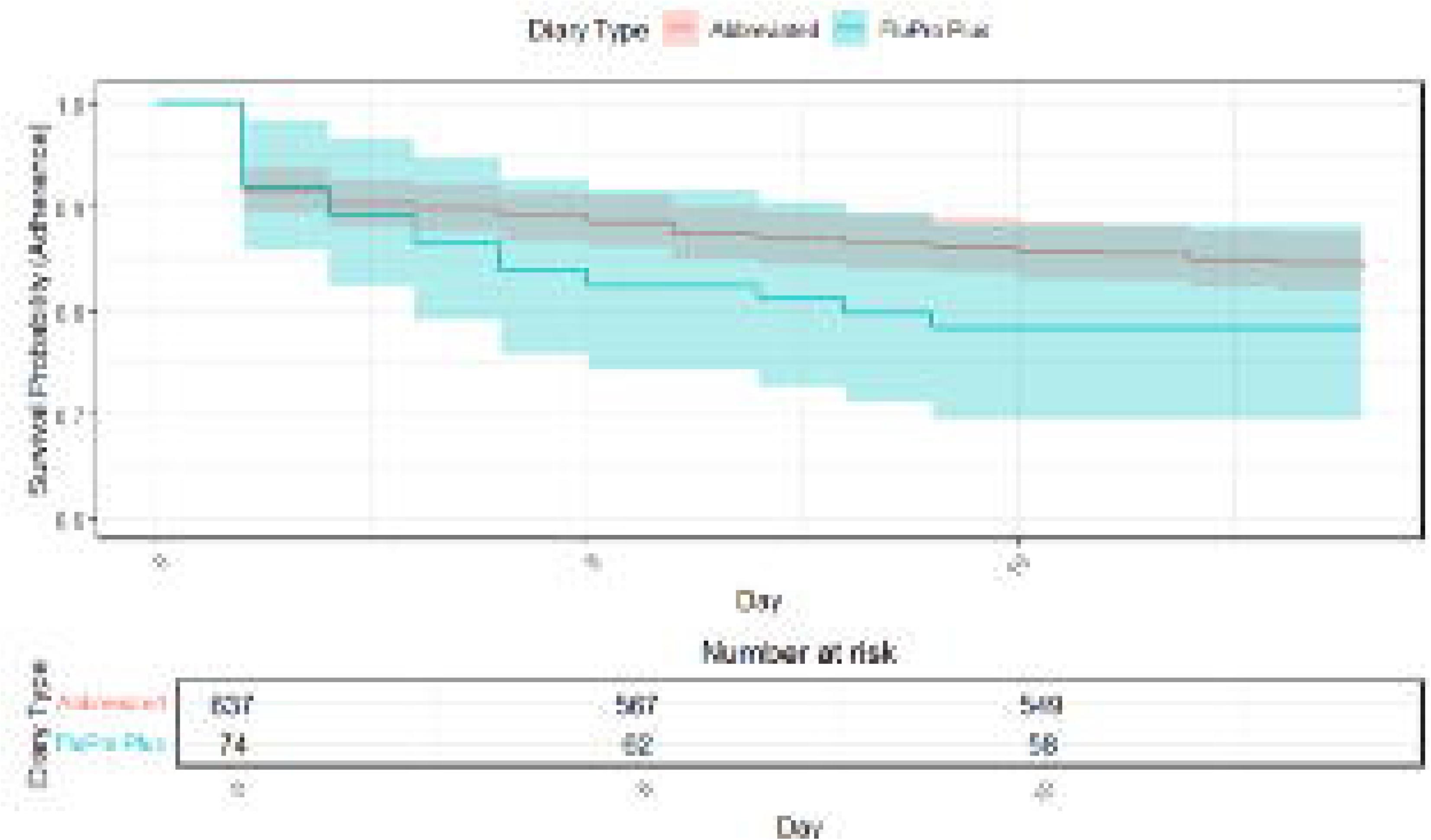
Kaplan Meier Survival Analysis Plot for Abbreviated and FluPro Plus diaries.

In the Cox model, dropout risk was higher among participants with severe symptoms (HR= 2.29, 95% CI: 1.31–4.00, p = 0.0037), and lower among those with better self-reported health (HR= 0.31, 95% CI: 0.14–0.69, p = 0.0039) and White participants (HR= 0.57, 95% CI: 0.35–0.93, p = 0.026). Diary type was not a statistically significant predictor but showed a trend toward higher dropout with the FLU-PRO Plus Diary (HR=1.78, 95% CI: 0.98–3.24, p = 0.057). A quadratic association between income and dropout was observed (HR= 1.98, 95% CI: 1.19–3.30, p = 0.008) **(Supplementary Table S2)**.

### Symptom trajectories and patterns

Across all participants, symptom severity declined significantly over the 14-day period (p < 0.001), with the largest reductions for muscle ache (β = –0.089), fatigue (β = –0.079), and difficulty concentrating (β = –0.066) **(Table 3**, **Figure 3)**. Diary type alone was not significantly associated with average symptom severity. Significant diary type x treatment interactions were observed for shortness of breath (p = 0.007) and muscle ache (p = 0.044). Three-way interactions (day × diary × treatment) were statistically significant for shortness of breath (β = 0.030, p < 0.001), muscle ache (β = –0.049, p < 0.001), and mood disturbance (β = –0.031, p < 0.001).

**Figure 3.**
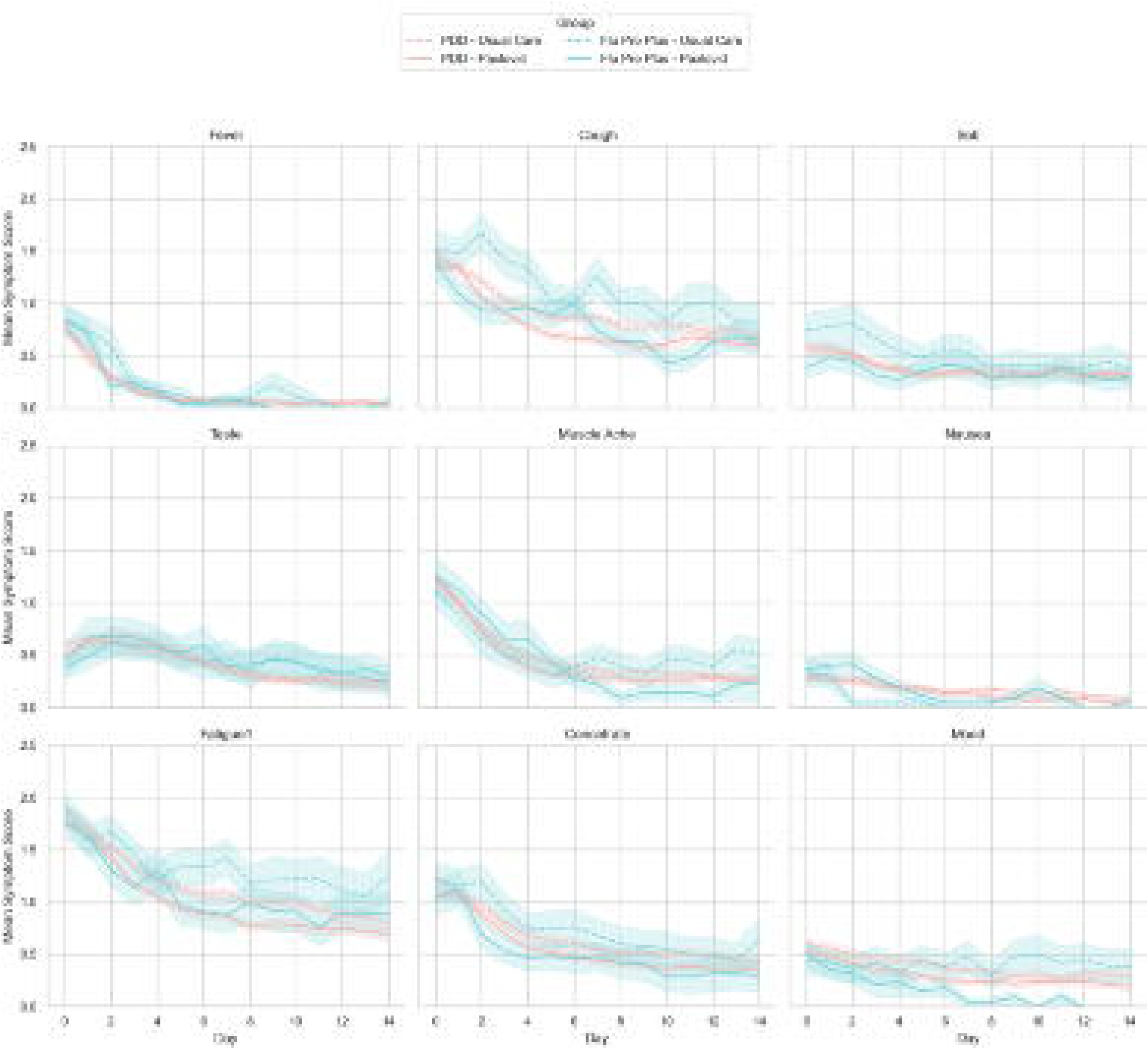
Symptom trajectories over 14 days for participants reporting daily symptoms via either the Abbreviated (PDD) or FluPro Plus (FPP) diary. Each subplot displays the mean severity score (0-3 scale) for one of nine common symptoms. Colored lines represent diary type (PDD in red, FPP in blue), and line style indicates treatment group (solid = Paxlovid, dashed, Usual Care). Shaded areas represent ± standard error of the mean (SEM).

**Table 3.**
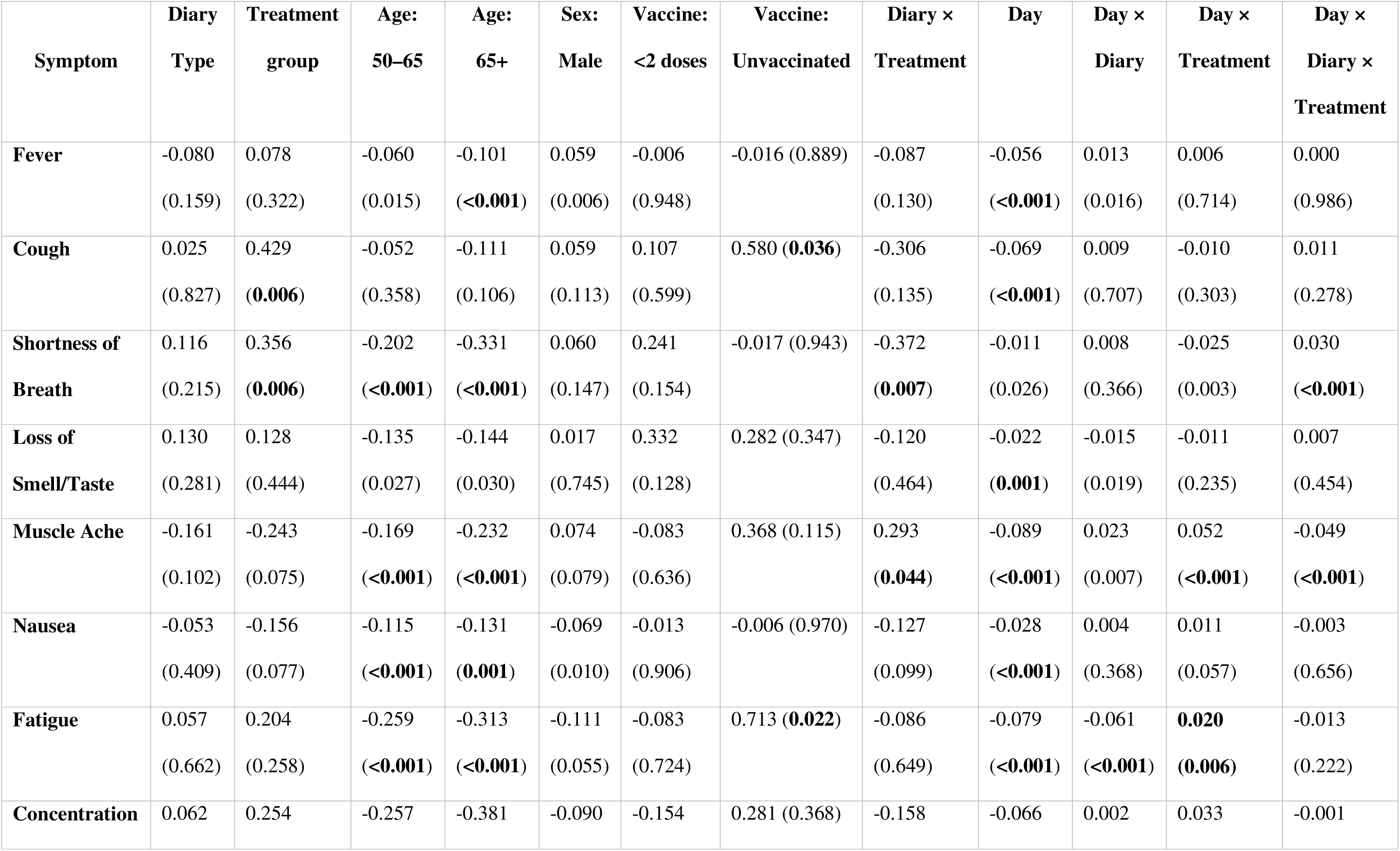
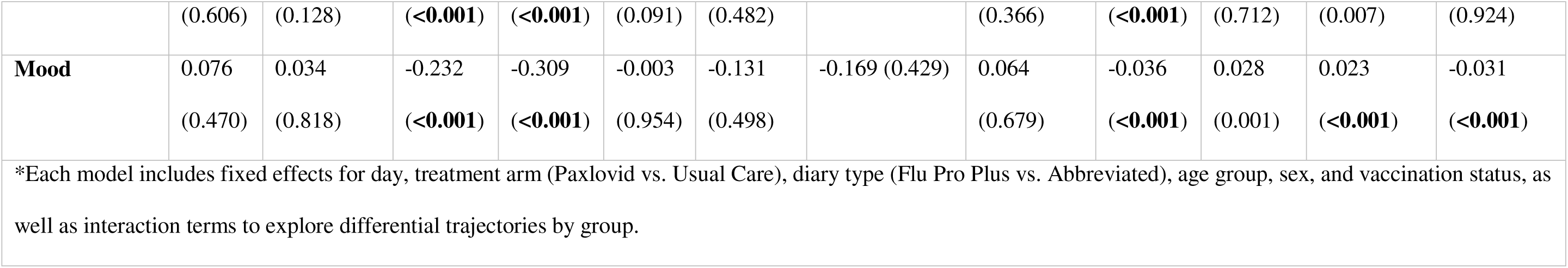
– Results (coefficient and p-value) of linear mixed-effects models assessing symptom severity trajectories over 14 days for nine common symptoms*.

Heatmaps showed similar symptom resolution patterns across groups, with faster improvement among Paxlovid recipients **(Supplementary Figure S1a-1d)**.

### Agreement between recovery metrics

#### Abbreviated Diary

In the Paxlovid group, agreement between self-reported recovery and return to usual health was strong (κ = 0.8371), with 2,104 person-days of concordant responses and 71 person-days where recovery was reported without a return to usual health. Agreement between recovery and return to usual activities was moderate (κ = 0.6278), with 627 person-days in which participants reported recovery but had not resumed their usual activities.

In the usual care group, agreement between recovery and return to usual health was also strong (κ = 0.8457), with 2,236 person-days of concordance. Agreement between recovery and return to usual activities was moderate (κ = 0.5772).

#### FLU-PRO Plus Diary

In the Paxlovid group, agreement between recovery and return to usual health was excellent (κ

= 0.8859), with only 6 discordant person-days out of 433 where recovery was reported. Agreement with return to usual activities was moderate (κ = 0.5273).

In the Usual Care group, agreement between recovery and return to usual health was strong (κ = 0.8512), and agreement with return to usual activities was moderate (κ = 0.6583), slightly higher than the corresponding value for the Paxlovid group.

### Predictive modeling of recovery using symptom diaries

In the Abbreviated Diary group, the GEE predictive models showed high discriminative performance: AUCs were 0.87 for recovery, 0.90 for return to usual health, and 0.87 for return to usual activities, with accuracies of 0.83, 0.86, and 0.77, respectively. Significant predictors across outcomes included fatigue, cough, shortness of breath, fever, nausea, muscle ache, taste disturbance, difficulty concentrating, and day.

In the FLU-PRO Plus group (common-symptoms model), AUCs were 0.92, 0.91, and 0.93, with accuracies of 0.88, 0.89, and 0.85, respectively. Predictors included cough, shortness of breath, fatigue, difficulty concentrating, mood, and day. Including all 34 symptoms modestly increased performance for recovery (AUC = 0.92, accuracy = 0.92) and return to usual activities (AUC = 0.94, accuracy = 0.88), with minimal change for return to usual health (AUC = 0.92, accuracy = 0.86) **(Figure 4)**.

**Figure 4.**
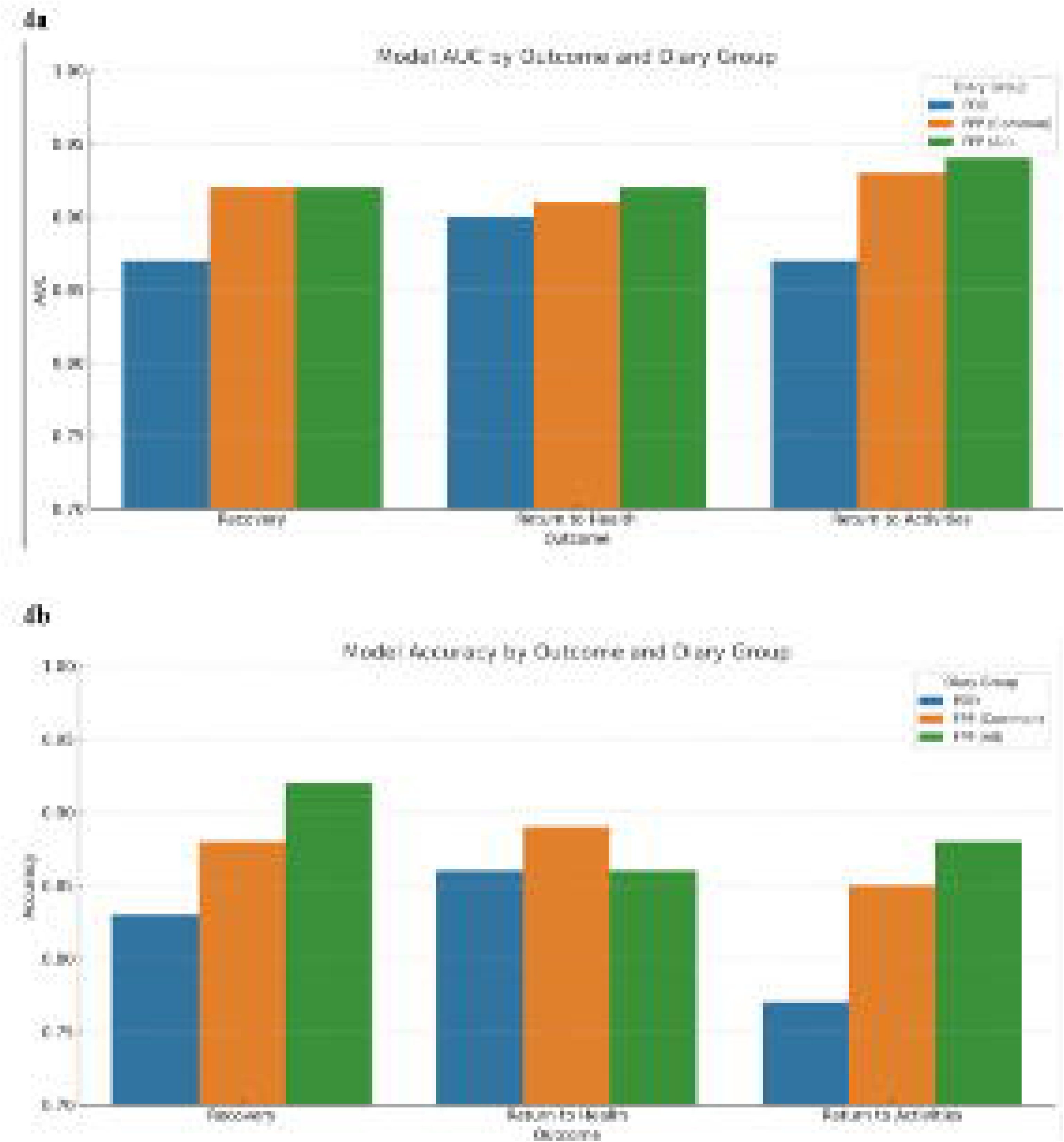
Predictive model performance for recovery outcomes across diary groups. Bar plots display the Area Under the Receiver Operating Characteristic Curve (AUC; 4a) and Accuracy (4b) for generalized estimating equation models predicting three recovery outcomes: “feeling recovered,” “return to usual health,” and “return to usual activities.” Models were stratified by diary group (Abbreviated (PDD) vs. FluPro Plus (FPP)), with the FPP group further divided into models based on common symptoms only and all available symptoms. All models accounted for repeated measures and included baseline and daily symptom predictors.

### Symptom predictors of time to recovery in the FLU-PRO Plus diary group

Of the 74 participants assigned to the FLU-PRO Plus diary, only those with complete daily entries and recovery status within the 14-day period were included in the Cox models. In the common-symptom Cox model (n = 56; 38 recovery events), baseline cough was significantly associated with delayed recovery (HR = 0.38, p < 0.005; C-index = 0.76). In the extended-symptom model (n = 53; 36 recovery events), cough remained significant (HR = 0.40, p = 0.02) with higher discrimination (C-index = 0.91). No other symptoms or demographic variables were statistically significant in either model.

## Discussion

In this secondary analysis of the CanTreatCOVID trial, we found that diary format, whether the abbreviated 9-item version or the longer 34-item FLU-PRO Plus, did not significantly affect either the likelihood of diary initiation or full 14-day compliance. However, treatment with Paxlovid was consistently associated with higher chances of diary completion and better overall adherence. Furthermore, while both diary formats demonstrated gradual declines in daily reporting, the reduction in response rates was slightly greater in the FLU-PRO Plus group, though not statistically significant.

Symptom severity declined over time across all groups, with notable interactions between diary type, treatment, and symptom trajectories such as shortness of breath, muscle ache, and mood disturbance. Agreement between participants feeling recovered and returning to usual health was strong to excellent, across both diary formats and treatment arms. Predictive performance for recovery-related outcomes was high for both diary types (AUC ∼0.87–0.94). Adding the extended FLU-PRO Plus items yielded only modest gains. Baseline cough was the only symptom that consistently predicted delayed recovery in time-to-event models.

Our findings are consistent with prior research on questionnaire length and adherence. Evidence from survey-methods literature indicates that shorter questionnaires are associated with higher completion rates without substantial losses in data quality^4–7^. In the context of patient-reported symptom diaries for respiratory infections, studies have reported mixed evidence on the added value of extended instruments compared to shorter formats. In a human influenza challenge study using the FLU-PRO diary, adherence was high (99%) and respondent burden low, but the extended format primarily confirmed responsiveness to change rather than offering clear superiority over shorter measures^12^. Similarly, evaluations of diary designs in other health contexts have shown that optimized, shorter diaries and standard, longer ones yield equivalent validity and acceptability, supporting the idea that concise formats can capture essential information without sacrificing data quality.^8,19^ In the context of SARS-CoV2, regulatory guidance emphasises balancing comprehensive symptom coverage with patient burden and recommends selecting key symptoms tailored to study objectives to minimise compliance challenges^20^. Our results align with this guidance, suggesting that a targeted symptom set, as used in the Abbreviated Diary, may be sufficient for tracking recovery trajectories while maintaining participant engagement.

Strengths of this study include the randomized, contemporaneous comparison of abbreviated and extended symptom diaries in a large outpatient cohort, conducted within the same trial framework and under identical operational conditions. Limitations should be considered when interpreting the results. The smaller sample size in the extended diary group may have reduced power to detect significant differences between formats; to mitigate this, we performed downsampling to create balanced comparison groups, which yielded consistent results. The 14-day follow-up period may not capture longer-term adherence patterns; while this timeframe reflects the acute phase of SARS-CoV-2 infection, future studies should include extended follow-up to assess longer-term engagement. Individual-level factors such as digital literacy, perceived burden, or patient preferences regarding diary format were not assessed, which limits insight into the drivers of engagement; this gap could be addressed in future mixed-methods evaluations. Finally, the trial population was predominantly White and middle-aged, which may constrain generalisability; subgroup analyses were conducted where possible, but more diverse recruitment is needed in future research.

## Conclusion

In this large outpatient SARS-CoV-2 trial, abbreviated and extended symptom diaries demonstrated comparable adherence, symptom trajectory patterns, and predictive performance for recovery-related outcomes. These findings support the feasibility of streamlined symptom monitoring tools in acute respiratory infection research and highlight opportunities for adaptive diary designs that balance comprehensiveness with usability. Future studies should evaluate diary performance over longer follow-up, in more diverse populations, and in settings with varying levels of digital access to ensure generalisability and scalability.

## Contributors

ADP, BH, PD, MG, PJ, TL, EM, SM, PS conceived of the work. KS managed coordination aspects of the project on a national level. DM and PB conducted the statistical analysis with inputs from BH and ADP. BH and ADP drafted the manuscript. All authors contributed to revising the manuscript for important intellectual content, gave final approval of the version to be published and agreed to be accountable for all aspects of the work.

## Data Availability

All data produced in the present study are available upon reasonable request to the authors.

## Acknowledgements

The authors thank our incredible patient and community partners: Brenda Andreas, Cris Carter, Jane Cooney, Gabriela Covaci, Letlotlo Gariba, Jennifer Hulme, Veronika Kiryanova, Kathy Kobow, Mike Lapenna, Mary Liu, Chris Maddison, Dorothy Nelson, Moon Ja Park, Lyric Paul, Donna Rubenstein, Dorothy Senior, Allard Schipper, Kimberly Strain, Margo Twohig, Mike Warren, John Zhan, and Alexander Zsager.

