## Supplementary File 1 for "Balancing data quality and participant burden: A comparative analysis of abbreviated vs extended symptom diaries in the CanTreatCOVID trial"

**Supplementary materials**

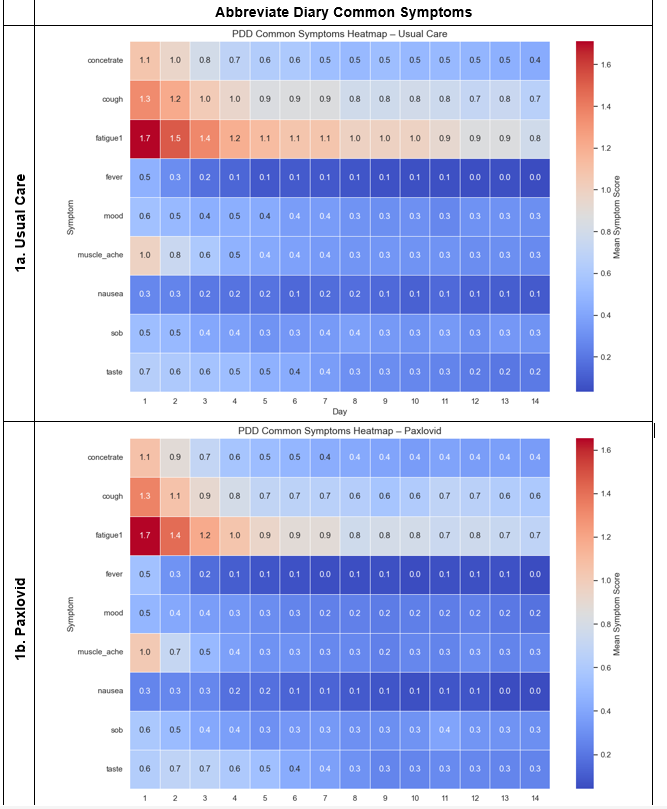

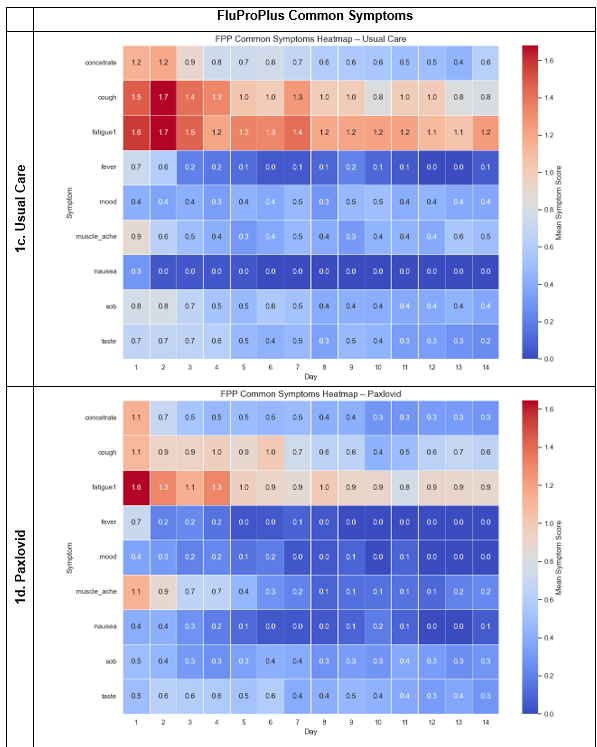

**Supplementary Figure S1- Symptom severity heatmap for common symptoms in the Abbreviated Diary (Panels 1a and 1b) and FLU-PRO Plus Diary (Panels 1c and 1d) groups, stratified by treatment group.**

Each heatmap illustrates the mean daily symptom severity scores from Day 1 to Day 14. Rows represent individual symptoms, and columns represent study days. Color intensity reflects the average symptom severity score on a 0–4 scale, with darker red indicating higher severity. Panels 1a and 1b correspond to participants using the Abbreviated Diary assigned to Usual Care and Paxlovid, respectively. Panels 1c and 1d show the same for the FLU-PRO Plus Diary group.

| **Supplementary Table S1- Diary completion rates by participant characteristics and diary type** | | | | |
| --- | --- | --- | --- | --- |
| **Variable** | **Group** | **FLU-PRO Plus** | **Abbreviated** | **p-value** |
| Treatment Group | Usual care | 58.59 | 63.31 | **0.011** |
|  | Paxlovid | 69.29 | 71.4 |  |
| Age | 18-49 | 64.54 | 65.16 | 0.113 |
|  | 50-65 | 66.66 | 69.29 |  |
|  | More than 65 | 54.98 | 65.38 |  |
| Sex | Female | 64.72 | 70.58 | 0.795 |
|  | Male | 68.13 | 69.83 |  |
| Race | Non-White | 41.85 | 53.53 | **0.002** |
|  | White | 69.55 | 71.19 |  |
| Education | High School or Less | 44.48 | 71.14 | 0.499 |
|  | University/College | 68.91 | 69.29 |  |
|  | Graduate | 61.12 | 71.72 |  |
| Housing | Renting/Other | 61.58 | 59.21 | 0.853 |
|  | Own Home | 65.45 | 70.73 |  |
| Employment | Non-Employed | 61.24 | 62.17 | 0.278 |
|  | Employed | 66.78 | 71.34 |  |
| Income | <60,000 | 70.89 | 67.10 | 0.349 |
|  | 60,000-100,000 | 58.52 | 71.76 |  |
|  | >100,000 | 64.57 | 70.83 |  |
| Chronic Disease | Yes | 68.96 | 70.37 | 0.102 |
|  | No | 52.27 | 69.72 |  |
| Self-reported health status | Poor/Fair | 72.14 | 67.11 | 0.444 |
|  | Good | 62.11 | 69.83 |  |
|  | Very good | 66.20 | 71.10 |  |
|  | Excellent | 64.71 | 72.11 |  |
| SARS-CoV2 symptom severity | No symptoms/Mild | 62.45 | 71.32 | 0.319 |
|  | Moderate | 66.04 | 70.53 |  |
|  | Severe/Very severe | 71.00 | 65.40 |  |

**Supplementary Table S2- Cox proportional hazards model results for factors associated with participant dropout**

| **Variable** | **Group** | **HR (95% CI)** | **p-value** |
| --- | --- | --- | --- |
| Diary Type | Abbreviated (ref) |  |  |
|  | FLU-PRO Plus | 1.78 (0.98 - 3.24) | 0.057 |
| Age | 18-49 (ref) |  |  |
|  | 50-65 | 0.64 (0.37 - 1.11) | 0.115 |
|  | More than 65 | 1.25 (0.63 - 2.48) | 0.515 |
| Sex | Male (ref) |  |  |
|  | Female | 0.89 (0.57 - 1.40) | 0.625 |
| **Race** | Non-White (ref) |  |  |
|  | White | **0.57 (0.35 - 0.93)** | **0.026** |
| Education | High School or Less (ref) |  |  |
|  | University/College | 1.36 (0.60 - 3.08) | 0.464 |
|  | Graduate | 1.22 (0.50 - 2.98) | 0.656 |
| Housing | Renting/Other (ref) |  |  |
|  | Own Home | 0.87 (0.50 - 1.51) | 0.622 |
| Employment | Non-Employed (ref) |  |  |
|  | Employed | 0.80 (0.49 - 1.31) | 0.38 |
| Income (Ordinal) | Linear Trend | 1.02 (0.68 - 1.53) | 0.912 |
|  | Quadratic Trend | **1.98 (1.19 - 3.30)** | **0.008** |
| Chronic disease | Yes (ref) |  |  |
|  | No | 1.42 (0.82 - 2.48) | 0.211 |
| Self-reported health status (Ordinal) | Linear Trend | **0.31 (0.14 - 0.69)** | **0.0039** |
|  | Quadratic Trend | 0.70 (0.38 - 1.28) | 0.248 |
|  | Cubic Trend | 1.21 (0.78 - 1.86) | 0.393 |
| SARS-CoV2 symptom severity (Ordinal) | Linear Trend | **2.29 (1.31 - 4.00)** | **0.0037** |
|  | Quadratic Trend | 0.87 (0.59 - 1.27) | 0.459 |
